# Pediatric pharmacogenomics from whole-exome sequencing: developmentally appropriate interpretation in 1,159 Russian children and newborns

**DOI:** 10.64898/2026.08.21.26360945

**Authors:** Anastasiia A. Buianova, Valery V. Cheranev, Mikhail Iu. Kuznetsov, Zhanna A. Repinskaia, Vera A. Belova

## Abstract

**Introduction:** The application of pharmacogenomics (PGx) in pediatrics is limited by the lack of age-oriented interpretation approaches, as algorithms developed for adults do not account for ontogenetic changes in the activity of drug-metabolizing enzymes and transport proteins. The aim of this study was to evaluate the clinical applicability of pharmacogenomic data in Russian children, assess the concordance between genotype-based recommendations and the ontogenetic status of drug-metabolizing enzymes, and develop recommendations for the generation of age-oriented PGx reports.

**Methods:** We analyzed whole-exome sequencing (WES) data from 524 pediatric patients and 635 newborns, filtering pharmacogenomic annotations according to PharmGKB/ClinPGx evidence levels (1A–2B) and the presence of the “Pediatrics” tag. The concordance between genotype-based recommendations and the ontogenetic status of drug-metabolizing enzymes was assessed in newborns. In a pediatric subgroup of 100 patients, a retrospective analysis of medical records was performed to evaluate the structure of pharmacotherapy and the frequency of adverse drug reactions (ADRs). A “PGx–ADR–cost” database was created, and the relative population burden index was calculated for 27 gene–variant–drug–ADR associations.

**Results:** Clinically relevant annotations (requiring drug avoidance or dose modification) accounted for only 5% of all initial pharmacogenomic annotations in both cohorts; 67.6% (pediatric cohort) and 67.2% (neonatal cohort) of these were related to alleles with altered function. Concordance between genotype-based recommendations and the ontogenetic status of drug-metabolizing enzymes in newborns was observed in only 5 of 14 (35.71%) gene–drug pairs. ADRs were identified in 21% of the 100 pediatric patients; however, only two cases could be explained by high-evidence PharmGKB/ClinPGx annotations. Ranking by relative population burden identified *UGT1A1*28*–irinotecan-induced neutropenia and *HLA-A*31:01*–carbamazepine-induced severe cutaneous reactions as priority associations.

**Conclusions:** Age represents a critical factor in the interpretation of pharmacogenomic data in children, as current approaches to PGx reporting do not adequately incorporate the ontogenetic context. We propose a pediatric PGx interpretation model that includes mandatory reporting of patient age, ontogenetic adjustment, evidence-level stratification, and multidisciplinary clinical assessment. Prospective validation is required to confirm the clinical utility of the proposed approach.

## 1. Introduction

The principle that “a child is not a small adult” is particularly relevant for pharmacogenomics (PGx), as the clinical interpretation of genetic variants depends not only on genotype but also on patient age, developmental stage, and specific characteristics of drug pharmacokinetics (PK) [1,2]. Unlike adults, children undergo dynamic ontogenetic changes in the activity of drug-metabolizing enzymes, transport systems, and elimination pathways, which may substantially influence the relationship between genotype and pharmacological phenotype [3–5].

However, current drug prescribing information generally recommends gradual dose adjustment based only on age, body surface area, or body weight. Recent studies demonstrate that age-dependent regulation of pharmacogene expression represents one of the major factors limiting the direct transfer of adult-derived PGx recommendations into pediatric clinical practice [6,7]. The most pronounced developmental changes have been described for cytochrome P450 (CYP)-dependent metabolic pathways, including *CYP3A4/3A7*, *CYP1A2*, *CYP2C19*, and *CYP2D6*, as well as for phase II metabolic enzymes and drug transporters [8]. Consequently, the same genetic variant may have different clinical implications in newborns, young children, and adolescents [9].

For example, age-related changes in *SLCO1B1* expression may modify the effect of genetic variants affecting transporter function on simvastatin exposure in children, emphasizing the need to integrate genotype with ontogenetic information [10]. Another clinically important example is the impact of *CYP2D6* variants on codeine metabolism, which has been associated with severe adverse drug reactions (ADRs) in children due to ultrarapid conversion of codeine to morphine [11,12]. Furthermore, codeine use is contraindicated in breastfeeding women [13]. Studies investigating drug excretion into breast milk remain limited due to ethical constraints and often rely on animal models. Moreover, the same medication may demonstrate teratogenic effects following transplacental exposure (e.g., hyperbilirubinemia in children exposed to atazanavir) [14], while being considered safe during breastfeeding. Azathioprine is an example of a medication considered compatible with breastfeeding [15]. Furthermore, carbamazepine monotherapy for epilepsy has even been associated with increased IQ scores in infants breastfed by treated mothers [16].

Despite the clear relevance of treatment personalization in pediatric populations, drug use in children continues to be characterized by substantial limitations in the evidence base. A considerable proportion of pediatric prescriptions are made outside approved age indications; according to the KiGGS study, approximately one-third of medications used in outpatient pediatric care in Germany had not been evaluated in the corresponding age group. Under these circumstances, PGx has the potential to improve treatment safety. According to Roberts et al., pharmacogenomic information may influence drug selection or dosing in approximately half of pediatric patients [17]. Furthermore, a large proportion of children receive medications for which Clinical Pharmacogenetics Implementation Consortium (CPIC) recommendations are already available. Wittwer et al. demonstrated that in Switzerland, 66.1% of children (159,172 individuals) received at least one medication associated with pharmacogenetic recommendations over a five-year period, with 96% of all potential gene–drug interactions attributable to only seven genes [18,19].

Nevertheless, implementation of PGx into pediatric practice faces not only biological but also interpretational challenges. Current PGx reports are often generated primarily based on identified genetic variants without comprehensive consideration of patient age, ontogenetic context, evidence level, or clinical applicability of specific recommendations, resulting in largely automated and decontextualized interpretation approaches [20]. Although ClinPGx contains a dedicated category of pediatric recommendations, the use of age-specific filters and restrictions is not mandatory in all existing approaches for generating PGx reports [21].

A particular challenge is presented by reports generated from whole-exome sequencing (WES) data, as this technology does not allow comprehensive assessment of several pharmacogenetically relevant features, including copy-number variation of *CYP2D6* [22]. In addition, inclusion of variants with limited clinical relevance may reduce confidence in PGx as a predictive medicine tool and bring such reports closer to consumer genetics approaches, where GWAS-derived associations are used to predict complex traits without established clinical utility [23].

Successful implementation of PGx requires not only genetic testing technologies but also a clinical infrastructure for interpretation [24,25]. In the Russian Federation, individual stages of the PGx workflow are distributed among clinical geneticists, laboratory geneticists, and clinical pharmacologists; however, a unified model of responsibility for translating genetic findings into clinical decisions is currently lacking. International experience demonstrates that PGx integration is most effective when supported by clinical decision-support systems incorporating evidence-based recommendations and automated treatment adjustment algorithms [26,27].

Thus, the key challenge in contemporary pediatric PGx is the transition from simple identification of genetic variants toward age-oriented clinical interpretation [28,29]. The aim of this study was to evaluate the characteristics of pharmacogenomic data application in children, assess the influence of ontogenetic factors on genotype–pharmacological phenotype concordance, and develop recommendations for the generation of standardized pediatric PGx reports.

## 2. Materials and Methods

### 2.1. Study design

The present study combined a retrospective PGx analysis of WES data from pediatric patients with a structured literature review of pharmacogene ontogeny. The study consisted of four complementary components: (i) identification of clinically relevant pharmacogenomic variants in neonatal and pediatric cohorts; (ii) retrospective analysis of prescribed pharmacotherapy in a subgroup of pediatric patients; (iii) integration of PGx annotations with published data on the healthcare costs associated with ADRs; and (iv) analysis of literature describing pharmacogene ontogeny to interpret the obtained findings from the perspective of age-dependent pharmacology.

The study was conducted in accordance with the principles of the Declaration of Helsinki and was approved by the Local Ethics Committee of the Pirogov Russian National Research Medical University (protocol No. 241, June 26, 2024). Written informed consent for the use of anonymized genomic data was obtained from all study participants or their legal representatives.

### 2.2. Study cohorts

WES data were obtained from an institutional database of the Pirogov Russian National Research Medical University, which included 6,102 samples sequenced between 2020 and 2025. This database served as the source population from which age-specific subgroups were selected for the present study. The overall characteristics of the cohort and the sequencing protocol used have been previously described in our earlier publication [30].

The present study included two age-specific cohorts: a neonatal cohort consisting of 635 full-term newborns (cord blood was collected into EDTA tubes after cutting the umbilical cord from the maternal segment) and a pediatric cohort comprising 524 patients aged from 3 months to 18 years. Both cohorts were analyzed using identical laboratory protocols, sequencing platforms, and bioinformatic pipelines.

Additionally, a subgroup of 100 patients was randomly selected from the pediatric cohort for retrospective medical record analysis aimed at evaluating the structure of prescribed pharmacotherapy and its potential pharmacogenomic relevance.

### 2.3. WES and bioinformatic analysis

WES was performed according to a standardized protocol described in detail previously [31]. Briefly, genomic DNA isolated from peripheral blood was used for library preparation with the MGIEasy Universal DNA Library Prep Set (MGI Tech, Shenzhen, China). Exonic sequence enrichment was performed using Agilent SureSelect Human All Exon capture kits (versions v6–v8). In the pediatric cohort, 37 samples were sequenced using the v6 kit, 91 samples using v7, and 396 samples using v8, whereas all 635 samples from the neonatal cohort were prepared using the v7 kit. Following enrichment, libraries were sequenced on the DNBSEQ-G400 platform (MGI Tech) using paired-end sequencing (PE100) with a target mean coverage depth of approximately 100×.

The resulting sequencing data underwent standard quality control procedures, alignment to the GRCh38 human reference genome, duplicate removal, and variant calling. Single-nucleotide variants (SNVs) and short insertions/deletions (indels) were identified using bcftools v1.18 [32], followed by annotation against the dbSNP build 156 database. Copy-number variation (CNV) analysis was performed for samples generated using compatible exome capture kits with CNVkit v0.9.8 [33], and interpretation of detected CNVs was performed using ClassifyCNV v1.1.1 [34]. *HLA* genotyping was performed from BAM files using HLA-HD (version 1.7.0) [35], followed by conversion of results into G-group nomenclature up to the second field resolution.

Star-allele assignment was performed using PAnno for the following genes: *CYP2B6, CYP2C19, CYP2C8, CYP2C9, CYP2D6, CYP3A4, CYP3A5, CYP4F2, DPYD, NUDT15, SLCO1B1, TPMT,* and *UGT1A1* [36]. The obtained results were additionally verified according to the current PharmVar nomenclature [37]. The *MT-RNR1* gene was excluded from analysis because the WES protocol used in this study did not include mitochondrial DNA capture.

### 2.4. Pharmacogenomic annotation and variant prioritization

PGx annotations were obtained from the PharmGKB/ClinPGx Clinical Annotations database [38] and included information on the gene, variant or star-allele, associated drug, phenotype, evidence level, PharmGKB/ClinPGx clinical annotation score, predicted allele functional effect, and supporting references. Allele frequencies from the PharmGKB/ClinPGx database were used for comparison with population data; this database aggregates allele frequency information from the 1000 Genomes Project Phase 3 (accessed January 5, 2026).

Annotations were sequentially filtered according to the following criteria: (i) type of pharmacogenomic annotation (star alleles/*HLA* or SNVs); (ii) presence of an annotation indicating pediatric clinical relevance; and (iii) PharmGKB/ClinPGx evidence level ranging from 1A to 2B. After generation of the final set of clinically relevant variants, they were further classified according to the predicted functional effect of the allele (normal, decreased, no, or increased function).

For genes with established interpretation algorithms, diplotypes were translated into predicted metabolic phenotypes according to CPIC guidelines [39] and phenotype interpretation tables proposed by Caudle et al. [40]. The resulting phenotypes were subsequently categorized according to the expected clinical recommendations: treatment modification, dose reduction, dose increase, or no requirement for therapy adjustment.

### 2.5. Retrospective analysis of medical records

To assess the potential clinical applicability of the identified PGx findings, a subgroup of 100 patients was randomly selected from the pediatric cohort for retrospective analysis of medical records. Clinical data were extracted from medical documentation, including demographic characteristics, primary diagnosis, prescribed pharmacotherapy, and medication history, including documented drug hypersensitivity reactions (allergic history) and other ADRs potentially associated with medication use (e.g., severe toxicity, clinical deterioration during therapy, or reactions potentially related to individual susceptibility or dosing characteristics).

Each prescribed medication, as well as drugs associated with previously documented ADRs, was compared with the individual PGx profile of each patient using the PharmGKB Clinical Annotations database, the FDA Table of Pharmacogenomic Biomarkers in Drug Labeling, and CPIC guidelines. If a corresponding PGx association was absent from all three sources, it was recorded as lacking published pharmacogenomic evidence at the time of analysis.

The analysis was descriptive in nature and aimed to evaluate the potential clinical relevance of identified gene–drug associations under real-world pediatric clinical practice conditions.

### 2.6. Three-component model for reproducible estimation of the frequency of pharmacogenomically relevant drug use in pediatrics

The proportion of pediatric patients receiving a given drug (*f_drug_*), was estimated using a multiplicative three-component model:

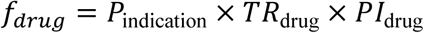

where: *P*_indication_ – the published prevalence of the target patient group (i.e., patients requiring a specific treatment) in the pediatric population, based on systematic reviews or registry data; *TR*_drug_ – the treatment rate, defined as the proportion of patients with a given indication who receive the specific drug, estimated from clinical guidelines, formulary data, or drug utilization studies; *PI*_drug_ – the persistence index, defined as the proportion of the disease period during which the patient actively receives the drug (calculated as the ratio of the mean treatment duration to the duration of the disease course). For drugs with multiple indications, contributions from all indications were summed:

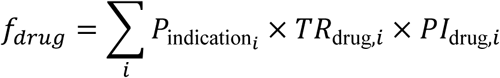

All parameters were extracted from published sources (Table S1). Where ranges of estimates were available, both lower and upper bounds were used to generate corresponding uncertainty intervals.

### 2.7. Development of an integrated “pharmacogenomics–ADR–cost” database for pediatric patients

An integrated database was developed by combining three independent layers of information for each “gene–variant–drug–ADR” combination included in the pediatric pharmacogenomic panel: (i) population allele frequency in the study cohort, (ii) PGx evidence describing the association between genotype and ADR risk, and (iii) published estimates of the cost associated with management of the corresponding ADR. Based on these data, an integrated relative burden score was calculated to prioritize pharmacogenomic variants according to their potential clinical and economic relevance.

PGx annotations, including gene, variant or star allele, associated drug, phenotype, ClinPGx clinical annotation score, evidence level, and CPIC functional allele classification (where available), were obtained from the ClinPGx Clinical Annotations database and restricted to annotations relevant to the pediatric population. Cost estimates for ADR management were collected through targeted searches of clinical and pharmacoeconomic literature. Allele and genotype counts were calculated based on the results of our pharmacogenomic analysis, which included 524 patients (1048 chromosomes).

For each ADR category, a representative treatment cost estimate (*cost_usd_mid*) was determined based on published data. When multiple estimates were available, the mean or the most representative value was selected based on literature analysis. Two ADR categories required specific methodological considerations. First, published cost estimates for cardiotoxicity reflected annual healthcare expenditures associated with patient management rather than the cost of a single ADR episode. Therefore, this parameter was explicitly classified as an annual cost and analyzed separately during visualization of the results. Second, because no unified published estimate was available for the cost of methotrexate toxicity management, a weighted average treatment cost was calculated by combining published data on glucarpidase-associated treatment costs with investigator-derived estimates of expenses associated with mild and moderate toxicity. For ADRs for which reliable published cost estimates could not be identified, corresponding values were not calculated and were not imputed.

For all combinations with available cost data, the relative burden score was calculated using the following formula:

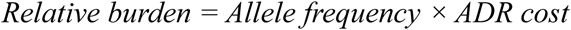

This metric was designed exclusively for comparative prioritization of variants within the investigated panel and reflects the combined contribution of risk-allele prevalence and estimated treatment costs of the associated ADR. It does not represent an estimate of expected healthcare expenditures, as it does not account for genetic variant penetrance, baseline ADR incidence, or population attributable risk.

It should be noted that several ADR cost estimates were derived from studies conducted in adult populations or were based on clinically comparable scenarios due to the lack of pediatric-specific data. Furthermore, calculations did not include inflation adjustment or differences in purchasing power between currencies. The relative burden score also does not incorporate information on genetic variant penetrance or baseline ADR incidence.

### 2.8. Literature review

To summarize current evidence regarding the ontogeny of proteins encoded by pharmacogenes, a structured literature search was performed in the PubMed/MEDLINE database. The search strategy was as follows: ((“pharmacogenetics”[Title/Abstract] OR pharmacogenomics[Title/Abstract]) AND (ontogeny[Title/Abstract] OR maturation[Title/Abstract] OR developmental[Title/Abstract]) AND (child*[Title/Abstract] OR pediatric*[Title/Abstract] OR neonatal[Title/Abstract] OR infant*[Title/Abstract])).

The search identified 122 publications published between 1995 and 2026. Titles and abstracts of all retrieved studies were screened for relevance to the research topic. Relevant publications underwent full-text review, focusing on studies addressing age-dependent expression of drug-metabolizing enzymes, drug transport proteins, pharmacogenomic biomarkers, and developmental modifications of genotype–phenotype relationships.

Particular attention was given to quantitative data describing ontogenetic trajectories of enzyme maturation, the age at which adult-equivalent functional activity is achieved, and clinical studies demonstrating the influence of developmental processes on the manifestation of pharmacogenetic effects.

## 3. Filtering of pharmacogenomic annotations and identification of actionable variants in neonatal and pediatric patients

We analyzed pharmacogenomic annotations in 524 pediatric patients (initially 3,724 annotations) and in an independent neonatal cohort (3,628 annotations). After sequential filtering according to variant type (star alleles/*HLA* versus SNVs), presence of the “Pediatrics” tag, and evidence level (1A–2B), 185 actionable annotations remained in the pediatric cohort (5% of the initial dataset) and 183 in the neonatal cohort (also 5%) (Figure 1A). Here and throughout the manuscript, the term “actionable” refers to pharmacogenomic annotations that are associated with alleles having functions other than normal (i.e., increased, decreased, no, or uncertain function), and therefore may require therapeutic adjustments such as dose modification, drug selection, or enhanced monitoring. The largest reduction occurred during filtering by the pediatric evidence tag: annotations lacking pediatric evidence were excluded (87.4% of star allele annotations and 86.2% of SNV annotations removed). Subsequent filtering by evidence level 1A–2B further reduced the number of annotations by 76.7–78.5%. Interestingly, the *UGT1A1*28* haplotype was present in 32.13% of newborns, with homozygous variants detected in 6.93% of individuals (associated with Gilbert syndrome).

**Figure 1.**
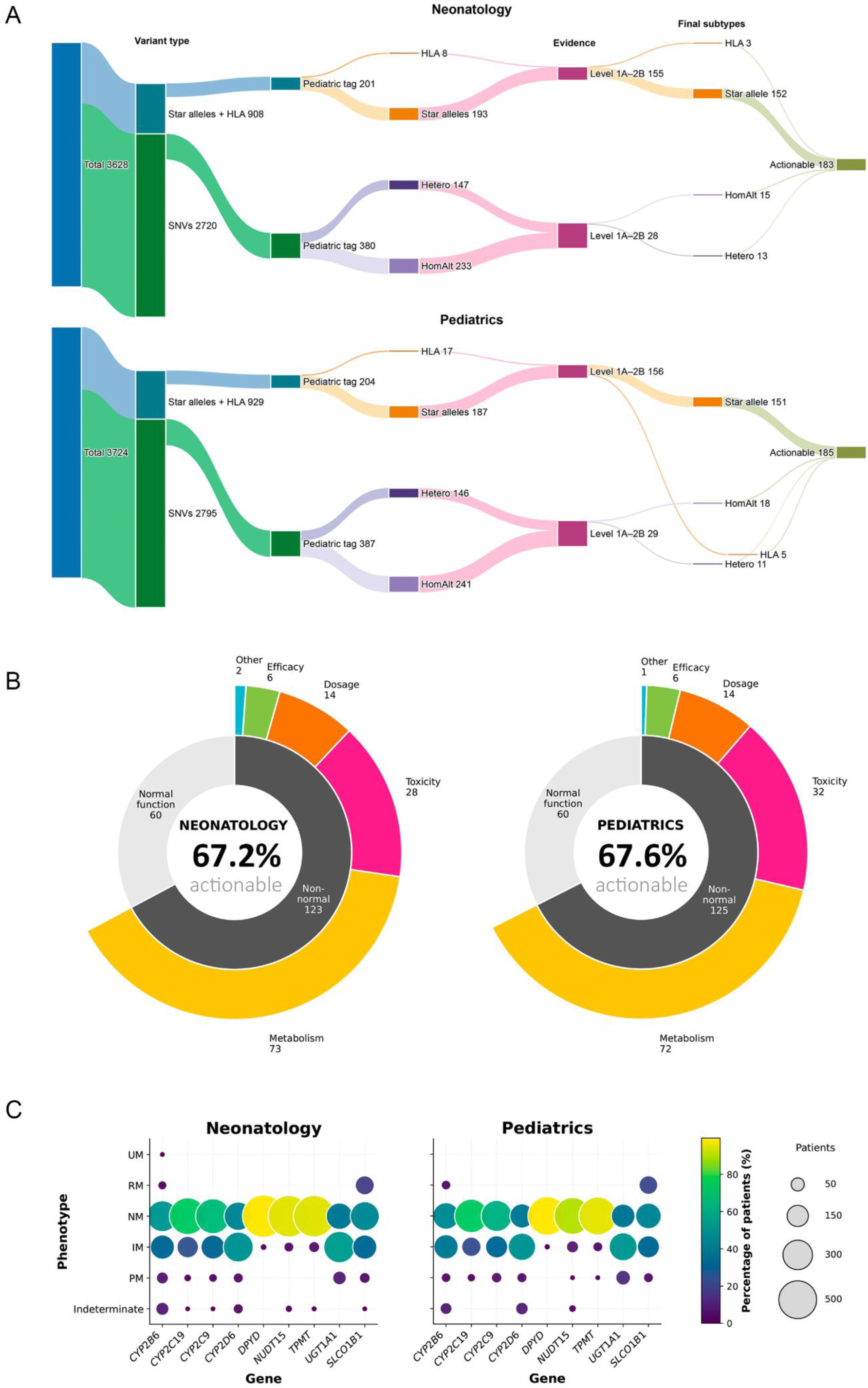
Distribution of pharmacogenomic annotations and metabolizer phenotypes in the study cohorts. (A) Workflow of pharmacogenomic annotation filtering in pediatric and neonatal cohorts. The Sankey diagram illustrates the stepwise reduction in the number of pharmacogenomic annotations after sequential application of filtering criteria. Node width is proportional to the number of annotations retained at each step. HomAlt – homozygous for the alternative allele. Actionable – associations involving alleles with functions differing from normal function. (B) Nested donut charts showing the distribution of pharmacogenomic annotations. The inner ring represents the proportion of alleles with normal function (light gray) and altered function, “actionable” (dark gray). The outer ring shows the distribution of altered-function alleles across phenotype categories (metabolism, toxicity, dosing, efficacy, and other). The center displays the total number of actionable annotations and the percentage of altered-function alleles. The predominance of metabolism-related annotations reflects the high contribution of cytochrome P450 enzyme polymorphisms in both cohorts. (C) Bubble heatmap showing the distribution of pharmacogenetic phenotypes across nine genes in pediatric and neonatal cohorts. Bubble size corresponds to the number of patients with a given phenotype. Bubble color indicates the proportion of patients with the phenotype within each cohort (scale shown on the right). Phenotypes are ordered according to functional relevance (top to bottom): UM – ultrarapid metabolizer; RM – rapid metabolizer; NM – normal metabolizer; IM – intermediate metabolizer; PM – poor metabolizer; Indeterminate. For consistency with other pharmacogenes, *SLCO1B1* phenotypes (normal function, increased function, decreased function, and poor function) were assigned to NM, RM, IM, and PM categories, respectively.

After excluding annotations associated with normal function (i.e., those not requiring therapeutic adjustment), 125 of 185 annotations remained in the pediatric cohort (67.6%) and 123 of 183 in the neonatal cohort (67.2%) (Figure 1B). The most frequent phenotype category was metabolism, accounting for 72/125 (57.6%) annotations in pediatrics and 73/123 (59.3%) in neonates, followed by toxicity (25.6% and 22.8%, respectively) and dosing (11.2% and 11.4%). The remaining annotations belonged to efficacy and “other” categories.

We mapped pharmacogenetic phenotypes for nine VIP genes that could be phased using PAnno (Figure 1C). Phenotype distributions were broadly similar between the pediatric and neonatal cohorts. *CYP2D6* showed a predominance of intermediate metabolizers (IM) (48.9% in pediatric and 48.7% in neonatal patients), followed by normal metabolizers (NM) (37.8% and 43.3%, respectively), whereas poor metabolizers (PM) accounted for 4.8% and 3.9%. Similar patterns were observed for *CYP2C19* and *CYP2C9*, with NM predominating (74.0–74.2% and 63.2–66.6%, respectively), while IM accounted for approximately 23–31% of individuals.

*UGT1A1* showed a substantial proportion of IM (51.1% in the pediatric cohort and 53.7% in the neonatal cohort), with PM accounting for 12.6% and 8.7%, respectively. For *SLCO1B1*, normal function was the most common phenotype (42.7% and 45.4%), followed by decreased function (31.3% and 31.2%), while poor function was observed in 4.6% and 4.9% of individuals.

For *TPMT* and *NUDT15*, NM predominated in both cohorts (>90%), whereas PM were rare (≤0.2%). *DPYD* showed an almost exclusively NM phenotype (>99%). Overall, phenotype frequencies were similar between pediatric and neonatal cohorts, although indeterminate calls were somewhat more frequent among pediatric patients, particularly for *CYP2D6* (8.6% vs. 4.1%) and *NUDT15* (1.9% vs. 1.3%).

## 4. Estimation of the frequency of pharmacogenomically relevant drug use and prioritization of “gene–variant–drug–ADR” pairs according to relative population burden

Figure 2A presents the estimated annual frequencies of use for 32 classes of pharmacogenomically relevant drugs in the general pediatric population (0–18 years). The most frequently prescribed drug classes were proton pump inhibitors (3%) and respiratory agents (salmeterol, 0.39%), followed by risperidone (0.29%), atomoxetine (0.16%), and oxcarbazepine (0.055%). In contrast, drugs with actionable pharmacogenomic recommendations but low frequency of use included warfarin (0.0053%), tacrolimus (0.0043%), and voriconazole (0.0037%). Oncology drugs and agents used for rare diseases (e.g., fluorouracil and irinotecan) demonstrated the lowest estimated frequencies, consistent with their limited use in routine pediatric practice.

**Figure 2.**
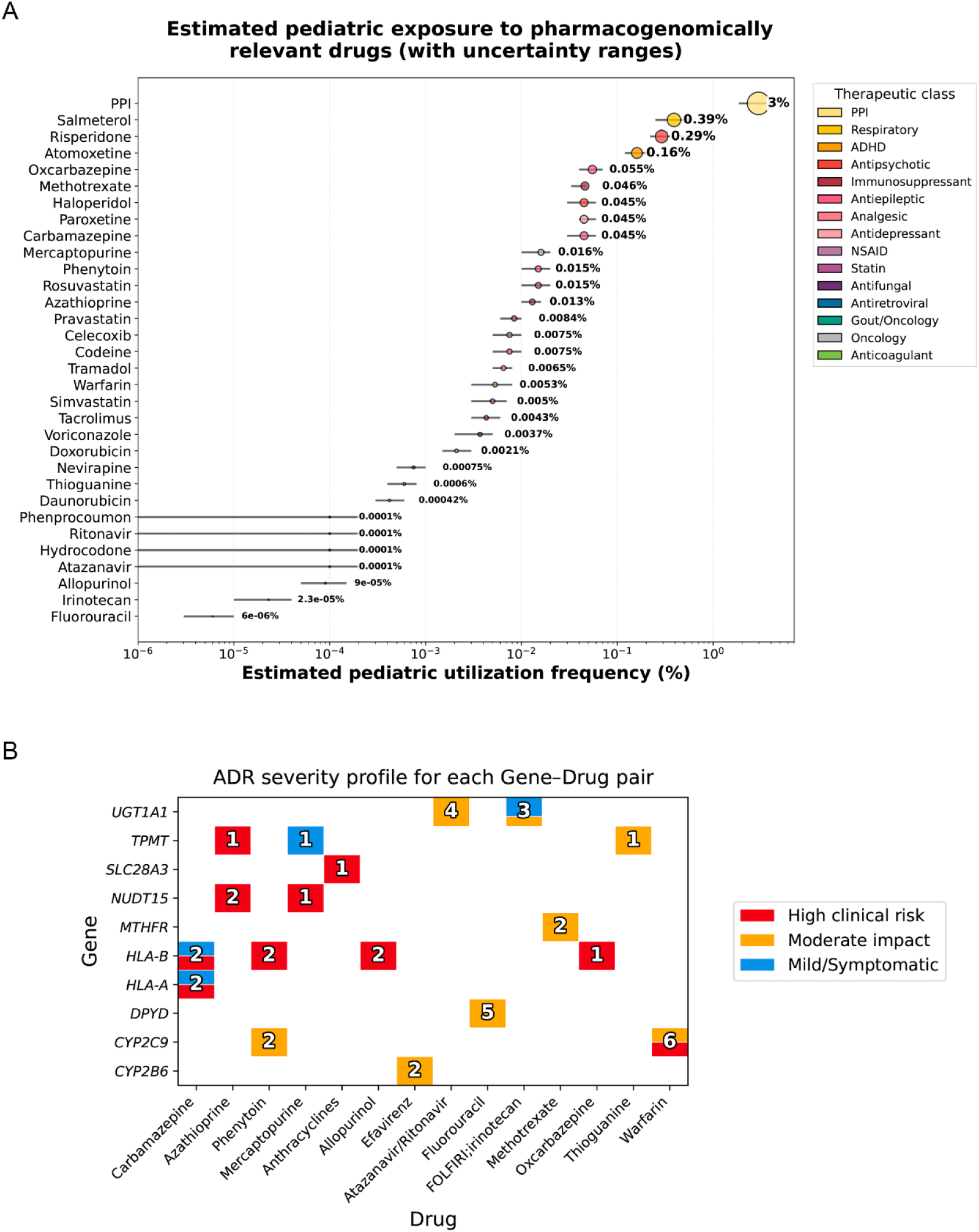
Estimated frequency of pharmacogenomically relevant drug use in children and severity of potential ADRs. (А) Bubble plot showing estimated frequencies of use for 32 pharmacogenomically relevant drug classes in the pediatric population (0–18 years). Bubble size corresponds to the median estimated frequency (%). Horizontal error bars represent the possible range (minimum and maximum estimates) based on published prescription data from heterogeneous cohorts (Table S1). For drugs with frequencies <0.01%, uncertainty ranges were expanded due to limited available data. (B) Heatmap showing the distribution of clinical severity of ADRs for each “gene–drug” pair with actionable pharmacogenomic annotations. Each cell is divided into colored segments proportional to the fraction of annotations associated with high (red), moderate (orange), and low (blue) clinical risk. The number within each cell represents the total number of unique annotations for the corresponding pair. Empty cells indicate the absence of annotations. Examples of high clinical risk include SJS/DRESS/TEN and bleeding events; moderate risk includes neutropenia and hyperbilirubinemia; low risk includes diarrhea and maculopapular rash.

The ADR profile matrix (Figure 2B) illustrates the clinical risk structure of “gene–drug” pairs based on pharmacogenomic annotations; the number within each cell indicates the total number of annotations for the respective pair. The highest number of unique annotations was observed for *DPYD*–fluorouracil (5 annotations: rs1801265, rs1801160, rs3918290 representing different genotypes), *UGT1A1*–atazanavir/ritonavir (4 annotations: *6, *28, *36, *37), and *CYP2C9*–warfarin (6 annotations: *2, *3, *11), reflecting the extensive evidence base supporting these interactions.

The highest clinical risk category (red) was observed for *HLA*-mediated reactions: all annotations involving *HLA-B* with carbamazepine and allopurinol were associated with SJS/TEN. The *CYP2C9*–warfarin pair included both high-risk outcomes (bleeding events) and moderate-risk outcomes (excessive anticoagulation), highlighting the importance of clinical context. For *UGT1A1*–irinotecan, the risk spectrum was heterogeneous, including moderate-risk neutropenia and low-risk diarrhea. In contrast, *TPMT*–mercaptopurine and *NUDT15*–azathioprine demonstrated exclusively high-risk myelosuppression outcomes despite a limited number of annotations (1–2 each). For *MTHFR*–methotrexate, no high-risk annotations were identified, suggesting a more favorable safety profile under standard dosing conditions.

We additionally calculated a relative population burden score (allele frequency × estimated ADR episode cost) for 27 “gene–variant–drug–ADR” pairs for which cost data were available (Figure 3; Table S2). The highest burden was observed for *UGT1A1*28*–irinotecan-induced neutropenia (relative burden = 5,253), driven by the high allele frequency (35%) and substantial treatment cost per episode ($15,000). This was followed by *CYP2C9*2/*3*–warfarin-induced bleeding (relative burden = 1,252 and 1,193; allele frequencies 10% and 9.5%, respectively; episode cost $12,500) and *HLA-A*31:01*–carbamazepine-induced SJS/DRESS/TEN (relative burden = 1,675; allele frequency 3.7%; episode cost $45,000).

**Figure 3.**
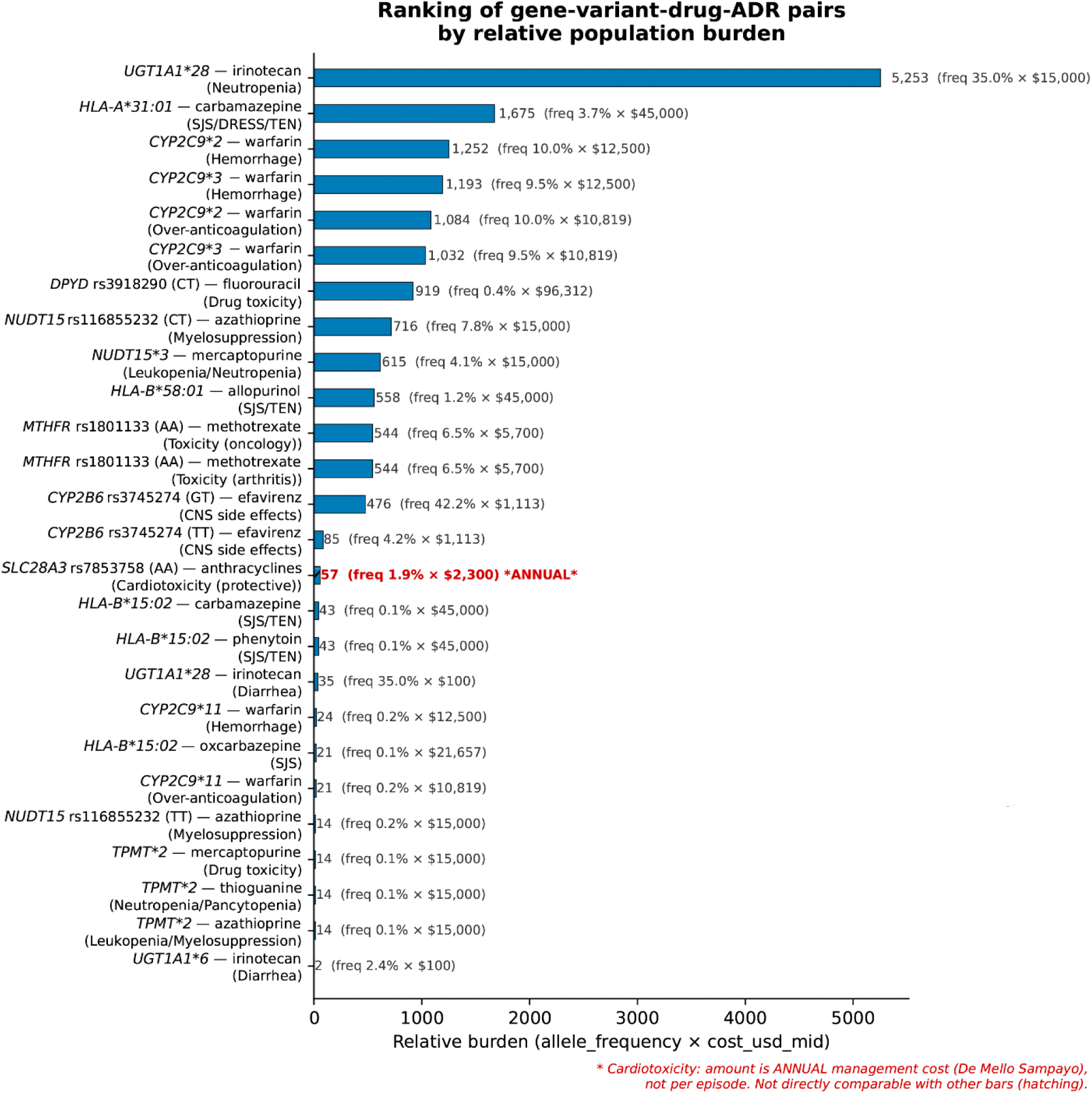
Ranking of gene–variant–drug–ADR pairs according to relative population burden. Bar plot ranking the relative population burden index (allele frequency × ADR episode cost) for 27 gene–variant–drug–ADR pairs with available cost estimates. The index is intended as a tool for relative prioritization rather than an estimate of absolute expected healthcare expenditure. Note: cardiotoxicity following anthracycline therapy corresponds to annual treatment costs.

## 5. Pharmacoeconomic Modeling: Assessment of the Potential Burden of Pharmacogenomically Associated Risks in the Pediatric Population

To illustrate the practical application of the pharmacoeconomic modeling framework, we selected two representative clinical scenarios with well-established pharmacogenomic associations and available epidemiological data: bronchial asthma (*ADRB2*–salmeterol) and postoperative anticoagulation after heart valve replacement (*CYP2C9*–warfarin). These examples were chosen to demonstrate the potential population-level impact of pharmacogenomic testing in pediatric practice.

The prevalence of bronchial asthma (BA) among Russian children in 2022 was 849.1 cases per 100,000 children [41]. A pharmacoeconomic study by A.Yu. Kulikov et al. demonstrated that combination therapy with salmeterol/fluticasone (commercial names Seretide® and Seretide® Multidisk®) was prescribed to 34.8% of patients with BA in the Russian Federation [42], although age-specific data were not provided.

In the present analysis based on the Russian pediatric cohort, the frequency of the ADRB2 rs1042713 AA genotype associated with reduced therapeutic response to salmeterol [PharmGKB, Tier 1 VIP evidence] was 12.2% (64 of 524 children). Therefore, among every 100,000 children in 2022, approximately 36 children could potentially have received salmeterol therapy despite carrying a genotype associated with reduced therapeutic response (assuming that 295.5 pediatric patients per 100,000 were treated with Seretide® or Seretide® Multidisk®).

Given a pediatric population of approximately 30 million individuals in the Russian Federation in 2022 [43], the estimated number of children with BA was approximately 254,730. Of these, approximately 88,643 were hypothetically treated with Seretide®/Seretide® Multidisk®, and 10,814 were estimated to carry the ADRB2 rs1042713 AA genotype. These patients could potentially experience reduced therapeutic efficacy, resulting in disease progression and increased healthcare utilization.

The cost of an emergency outpatient visit due to clinical deterioration was assumed to be 1,727.1 RUB, and the cost of an emergency medical service call was 3,288.9 RUB, according to the Government Resolution of the Russian Federation No. 2497 dated December 29, 2022, “On the Program of State Guarantees of Free Medical Care Provision for Citizens for 2023 and the Planning Period of 2024–2025.” Ultimately, this could lead to hospitalization, with the baseline cost of inpatient care in a 24-hour hospital setting estimated at 41,858.1 RUB. The total potential healthcare burden associated solely with suboptimal BA therapy was estimated at 452,653,493.4 RUB.

According to L.A. Bokeria et al., 894 pediatric heart valve replacement procedures were performed at the A.N. Bakulev National Medical Research Center of Cardiovascular Surgery of the Ministry of Health of the Russian Federation between January 1998 and January 2013 [44]. Therefore, the average annual number of procedures at this center was approximately 59.6 interventions per year. Following mechanical valve implantation, current clinical guidelines recommend lifelong anticoagulation therapy with vitamin K antagonists (warfarin) [45]. As a scenario assumption in the present analysis, all patients undergoing valve replacement were considered to require long-term warfarin therapy.

Based on genotyping data from the Russian pediatric cohort, the frequency of carriers of at least one *CYP2C9*2/*3* reduced-function allele was 182/524 ≈ 0.347 (34.7%). Therefore, the estimated number of patients receiving warfarin and carrying a genetic variant associated with reduced drug clearance was: 59.6 × 0.347 ≈ 20.7 ≈ 21 children per year.

Pharmacoepidemiological studies indicate that patients carrying reduced-function *CYP2C9* variants have a substantially increased risk of hemorrhagic complications during warfarin therapy. Assuming that severe hemorrhagic complications occur in only 2–3% of these patients, the expected incidence would be 0.42–0.63 cases/year. The cost of treating one major hemorrhagic complication (e.g., gastrointestinal bleeding requiring intervention and surgical management) in an inpatient setting was estimated at 477,600 RUB per case, based on real-world clinical data from City Clinical Hospital No. 15 named after O.M. Filatov, Moscow Department of Health (April 1, 2026). Therefore, annual costs associated only with treatment of severe bleeding events at a single center could reach approximately 191,040–286,560 RUB/year.

However, this estimate is substantially conservative because it does not account for:

1. The need for intensive warfarin dose optimization in each patient, including more frequent physician visits, additional laboratory monitoring (INR measurements), and dose adjustments.
2. Increased healthcare system burden, including additional consultations, laboratory testing, telephone consultations, and the risk of adverse outcomes associated with unstable anticoagulation.

In a study by Kim et al. [46], the mean cost of managing a single episode of over-anticoagulation or major bleeding in adults receiving warfarin was 10,819 ± 11,536 USD per treatment course. In pediatric populations, available data primarily concern hemophilia, with hospitalization costs for severe bleeding estimated at 10,000–50,000 USD [47]. Despite differences in bleeding etiology, these values provide an approximate estimate of the potential economic burden of hemorrhagic complications in pediatric practice because data specifically describing warfarin-induced ADRs in children remain limited.

## 6. Neonatal Pharmacogenetic Dosing Recommendations and Enzyme Ontogeny

In a separate analysis of 13 drugs approved for neonatal use (and an additional six drugs approved from 1–3 months of age), genotype-based dosing recommendations (ClinPGx) were compared with the actual ontogenetic status of drug-metabolizing enzymes during the first 28 days of life (Figure 4).

**Figure 4.**
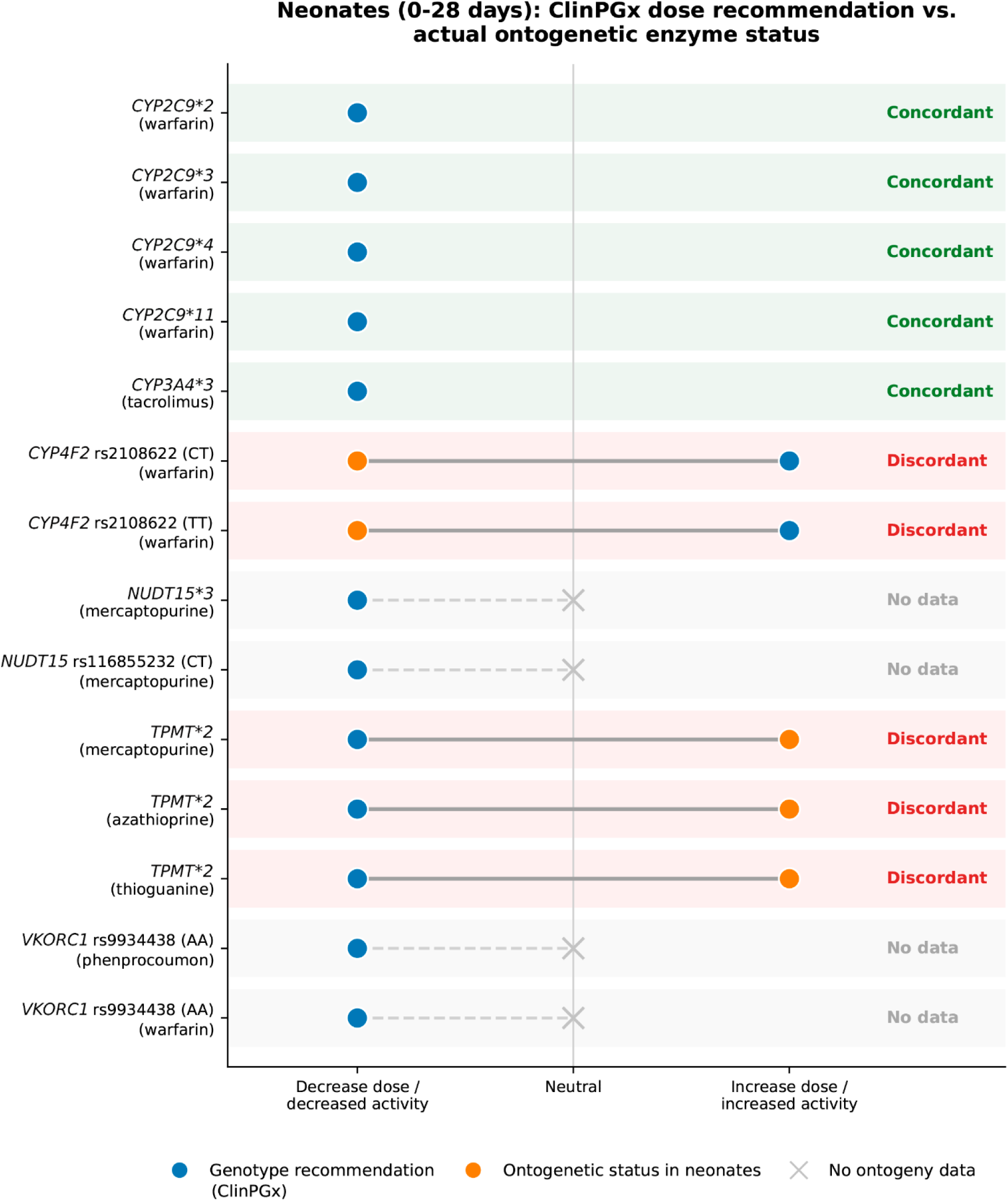
Neonatal dosing recommendations integrating pharmacogenomics and enzyme ontogeny. Gantt-like diagram comparing genotype-based dosing recommendations (ClinPGx, blue circles) with actual ontogenetic enzyme status in neonates (orange circles). Background colors indicate: green, concordance (recommendation consistent with ontogeny); red, discordance; gray, absence of available literature data.

Among the 14 analyzed gene–drug pairs, five (35.71%) demonstrated concordance (green background). For example, reduced-function *CYP2C9* variants (*2, *3, *4, 11) are associated with recommendations for dose reduction, which is consistent with CYP2C9 activity in neonates (Table 1), reaching approximately 30% of adult levels. *CYP3A4*3* is associated with tacrolimus dose reduction recommendations, consistent with low CYP3A4 activity at birth. Discordant results (red background) were observed for *CYP4F2* rs2108622 (CT/TT) with warfarin: the genotype-based recommendation suggests dose increase, whereas neonates have physiological vitamin K deficiency, potentially requiring dose reduction. *TPMT*2* with thiopurines was also discordant: the genotype suggests dose reduction, while TPMT activity in neonates is more than 50% higher, potentially indicating a need for higher doses. Ontogenetic data were unavailable for *NUDT15* and *VKORC1*.

**Table 1.** Ontogenetic maturation of clinically relevant pharmacogenes and its implications for pediatric PGx.

| Gene | Approximate maturation period | Genotypes where ontogeny is most critical | PGx phenotypes most affected by age | Clinical relevance | Applicability of adult PGx recommendations | Approximate maturation period |
| --- | --- | --- | --- | --- | --- | --- |
| *CYP3A7* → *CYP3A4* | Developmental switch from fetal to adult isoform; CYP3A7 dominant in fetal liver, CYP3A4 appears in first week of life | First 6–12 months of life | All genotypes (CYP3A4 expression is negligible at birth regardless of genotype) | All CYP3A4-related phenotypes | Midazolam, tacrolimus, cyclosporine metabolism; reduced CYP3A4 activity at birth → higher dose requirements in young children [5, 48, 49, 50] | Limited during neonatal period; CYP3A7-mediated metabolism differs substantially from adult CYP3A4 |
| *CYP2D6* | Genotype–phenotype relationships appear early in life; enzymatic capacity continues to increase during childhood | ~3–5 years to adult-like activity; phenotype correlates with genotype from early infancy [51] | *4, *5, *10, *17 (reduced/no function); gene duplications (UM) | UM, IM, PM phenotypes | Codeine (toxicity in UM via breastfeeding), tramadol, atomoxetine, antidepressants; UM mothers may cause neonatal respiratory depression [49, 50, 52, 53] | Cautious; genotype–phenotype correlation appears early but maturation continues; UM risk is amplified in neonates |
| *CYP1A2* | Absent in fetal liver; appears after birth; activity increases during infancy and may exceed adult levels during early childhood [48, 49, 51] | Progressive maturation through infancy; adult levels reached by 4–6 months (contested); peak > adult at 1–2 years [49, 53] | All genotypes (loss-of-function is exacerbated by low neonatal expression) | PM phenotypes (loss-of-function is amplified by ontogeny) | Caffeine, theophylline metabolism; higher clearance in children → higher weight-based doses [5, 48, 49, 53] | Not applicable in neonates; pediatric clearance may exceed adult |
| *CYP2C19* | 12–15% adult level at 8 weeks gestation; linear increase in first 5 months; adult levels by ~10 years; ontogeny may mask genotype–phenotype relationships in infants <4 months [54] | First 5 months (critical window), adult by 10 years | *2, *3 (no function); *17 (ultrarapid) | IM and PM phenotypes; reduced expression may attenuate group differences in early infancy | Omeprazole, pantoprazole, voriconazole; substantially different exposure in neonates; higher dose may be needed in rapid metabolizers [49, 50, 52, 53] | Not applicable before ~6 months; genotype effect is partially masked by ontogeny in early infancy |
| *CYP2C9* | 1–2% of adult in first trimester; ~30% at term; 35-fold interindividual variability in first 5 months [54] | ~1–2 years (adult level in 51% of samples by 5 months) | *2, *3 (reduced/no function); *5 (African-specific) | PM phenotypes | Phenytoin, warfarin, NSAIDs; higher weight-based phenytoin dose in children [48, 49, 52, 53] | Limited in early infancy; genotype effect becomes relevant as expression matures |
| *CYP2E1* | Appears in second trimester, sharply rises in third trimester and after birth; levels: 0.35 (II trim), 6.7 (III trim), 8.8 (neonate), 23.8 (30–90 d), 41.4 (>90 d–18 y) [55] | ~3 months to half of adult level; adult by ~18 years | All genotypes (low neonatal expression) | PM phenotypes (low baseline activity) | Acetaminophen (hepatotoxicity risk), halothane, ethanol; low neonatal CYP2E1 may protect from acetaminophen toxicity [5, 53] | Limited in neonates; metabolic activation pathways are immature |
| *UGT1A1* | Absent in fetal liver; increases after birth; adult level by 6 months; neonates: glucuronidation/sulfation ratio 0.34 vs. 1.8 in adults [49] | ~3–6 months | *28/*28 (reduced function); *6 (Asian-specific) | *28/*28 and other reduced-function alleles | Neonatal hyperbilirubinemia; irinotecan toxicity risk; glucuronidation capacity is reduced in neonates [5, 49, 52] | Not applicable in early infancy; adult levels by 6 months |
| *SULT1A1* | Relatively stable postnatal activity; may exceed adult in infants; sulfation matures faster than glucuronidation [51, 56] | From birth (matures faster than UGT) | Functional variants (e.g., *2) | All functional variants | Acetaminophen metabolism (compensates for low UGT activity in neonates); conjugation reactions [5, 49, 51] | Potentially applicable from birth; sulfation is mature earlier than glucuronidation |
| *NAT2* | Absent until 11–14 weeks; appears by 16 weeks; all neonates are phenotypically slow acetylators until 55 days [51] | Neonatal slow phenotype until 55 days; adult phenotype by 3–4 years | *4 (rapid), *5, *6, *7 (slow) | Slow acetylator phenotype (all neonates are phenotypically slow regardless of genotype) | Isoniazid, procainamide, dapsone, sulfasalazine; genotyping is not informative in first 55 days [48, 49, 52] | Not applicable in first 55 days of life; genotype masked by ontogeny |
| *TPMT* | Relatively stable from birth; erythrocyte activity may be 50% higher in neonates; distribution of phenotypes same as adults [51] | Present from neonatal period | *2, *3A, *3C (no function) | Intermediate and poor metabolizers | Thiopurine-induced myelotoxicity (mercaptopurine, azathioprine); dose reduction 8–15-fold in poor metabolizers [49, 50, 52] | Applicable from birth; genotype–phenotype correlation is stable |
| *CYP3A5* | Genotype-driven expression established early; polymorphically expressed in ~25% of adults [49] | From birth | *1/*1, *1/*3 (expressors); *3/*3 (non-expressors) | Expressor vs. non-expressor phenotypes | Tacrolimus dosing; African Americans have higher frequency of *1 allele → higher dose requirements [5, 50, 52] | Applicable with caution; genotype effect is present from birth but may be partially masked by CYP3A7 in early infancy |
| *NUDT15* | No data | From birth | *2, *3 (no function); rs116855232 (CT, TT) | Intermediate and poor metabolizers | Thiopurine toxicity (mercaptopurine, azathioprine, thioguanine); higher risk in Asian populations [57] | Applicable from birth |
| *SLCO1B1* | Age-dependent expression; conflicting data in children; pediatric studies show opposite effect to adults [58] | Not clearly defined — requires further pediatric studies | *5 (rs4149056); *15 | Reduced-function phenotypes; effect direction may differ in children | Statins, methotrexate disposition; pediatric studies show lower concentrations in variant carriers (opposite to adults) [52] | Limited data; adult PGx recommendations may not apply to children; direction of effect may be reversed |
| *ABCB1*<br>(P-gp) | Enterocyte expression increases rapidly 3–6 months, adult levels by ~2 years; blood-brain barrier expression ↑ with age [52] | Enterocyte P-gp: ~2 years; CNS: increases with age | C3435T, G2677T/A (affect expression/function) | All phenotypes (lower P-gp in neonates may increase CNS toxicity) | CNS drug penetration, intestinal absorption; immature P-gp may increase risk of CNS toxicity in neonates [5, 51, 52] | Cautious; P-gp expression is age-dependent, affecting drug distribution |
| *VKORC1* | Age is more important than genotype in children; age explains 28.3% of dose variation vs. 3.7% for *VKORC1* [59] | Children — age dominates VKA dosing until ~20 years | -1639G>A (AA, GA, GG) | All genotypes (age dominates dosing) | Warfarin, phenprocoumon dosing; age, not genotype, is primary determinant in children; adult algorithms underestimate pediatric dose [59] | Not applicable; age dominates genotype in pediatric VKA dosing; adult algorithms underestimate pediatric dose |
| *CYP4F2* | Lowest at birth (0–12 days); increases in later age groups [60] | Neonates/infants — low *CYP4F2* activity may affect warfarin dosing | V433M | Age-dependent expression; neonatal vitamin K physiology may influence warfarin response independently of genotype [60] | Warfarin dosing; *CYP4F2* polymorphisms do not significantly affect warfarin maintenance dose in children [61] | Limited in neonates; physiological factors may dominate |
| *DPYD* | No ontogeny; fetal/neonatal expression confirmed by congenital DPD deficiency cases [62] | From birth (genotype determines risk) | rs3918290 (c.1905+1G>A, *2A); others | Poor metabolizers (dihydropyrimidine dehydrogenase deficiency) | Fluorouracil, capecitabine toxicity; severe toxicity in carriers of loss-of-function variants [50] | Applicable from birth for risk assessment; testing recommended before fluoropyrimidine therapy |
| *HLA-B* | Genotype-dependent; no ontogenetic effect | From birth | *15:02, *58:01, *31:01, *57:01 | Presence/absence of risk allele | Carbamazepine, allopurinol, oxcarbazepine, phenytoin; SJS/TEN; DRESS syndrome [PharmGKB] | Applicable from birth; *HLA* genotype is stable throughout life |
| *MTHFR* | Genotype primarily; conflicting evidence; no clear ontogeny | No clear ontogeny; phenotype may be age-independent | rs1801133 (C677T; AA, AG, GG) | All genotypes | Methotrexate toxicity (conflicting evidence); low clinical actionability (DPWG “no recommendation”) [PharmGKB] | Not clinically actionable; CPIC/DPWG do not recommend routine testing |

This is well illustrated by a real-life clinical study by Rodieux et al. [9], which evaluated *CYP450* genotyping and phenotyping in 48 children (mean age 9.7 years). The concordance between predicted (genotype-based) and measured (probe-based) phenotypes was highly variable: 80% for *CYP2C9*, 50% for *CYP2D6*, 37.5% for *CYP2C19*, and only 33% for *CYP3A5*. In about half of the non-concordant cases for *CYP2C19* and *CYP3A5*, the discrepancy was explained by drug-induced phenoconversion, while for *CYP2D6*, the poor detection of ultrarapid metabolizer alleles contributed to the mismatch. Importantly, genotyping or phenotyping results explained or contributed to the clinical event (ADR, inefficacy, or low plasma concentration) in 56% of cases.

## 7. Analysis of Drug Therapy and Adverse Drug Reactions Based on Medical Records in the Pediatric Cohort

Analysis of medication histories from 100 patients demonstrated that the most frequently prescribed drug classes were antibiotics (68%), glucocorticosteroids (50%), neuroprotective agents (31%), vitamins (27%), and immunosuppressants (23%) (Figure 5A). Among antibiotics, penicillins (18% of patients) and cephalosporins (15%) predominated. The top five most frequently prescribed individual drugs included amoxicillin/clavulanate (18%), prednisolone (16%), ceftriaxone (12%), dexamethasone (10%), and meropenem (8%) (Figure 5B). The highest proportion of treatment inefficacy was observed for biological agents: infliximab was ineffective in 60% of patients (3/5), and adalimumab in 50% (1/2). Among anticonvulsants, valproic acid showed no clinical effect in 37.5% of patients (3/8), while levetiracetam was ineffective in 33.3% (2/6) (Figure 5C).

**Figure 5.**
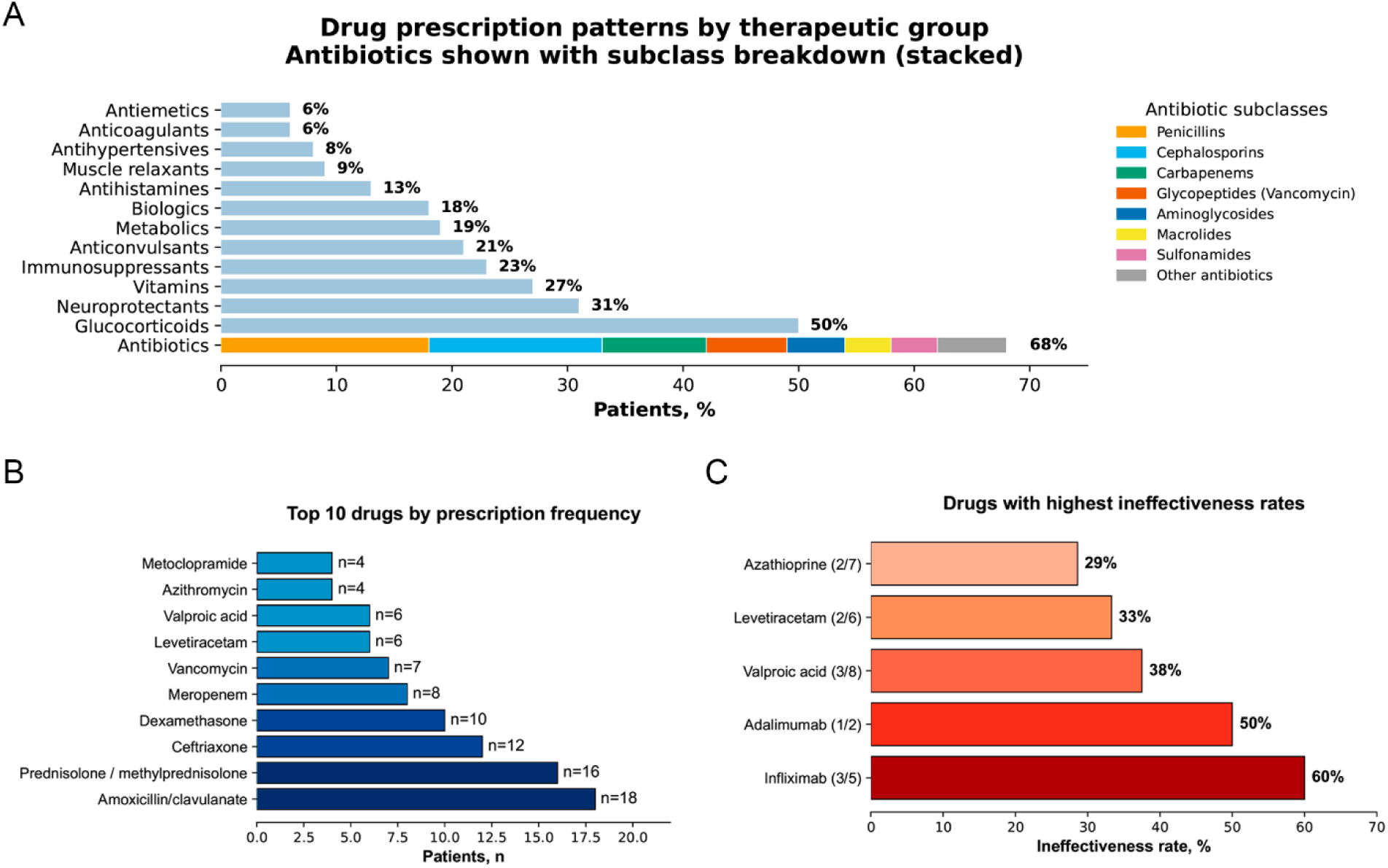
Structure of prescribed therapies and treatment inefficacy in the pediatric cohort (*n* = 100). (A) Stacked bar chart showing therapeutic drug classes; antibiotics (68%) are further subdivided by subclasses, with penicillins (*n* = 18) and cephalosporins (*n* = 15) being predominant. (B) Top 10 individual drugs by prescription frequency; the most frequently prescribed drugs were amoxicillin/clavulanate (*n* = 18) and prednisolone/methylprednisolone (*n* = 16). (C) Drugs with the highest proportion of treatment inefficacy; the highest rates were observed for infliximab (60%; 3/5) and adalimumab (50%; 1/2).

ADRs were identified in 21% of patients. The majority were associated with antibacterial therapy: allergic reactions to penicillins were documented in 6 patients (in 2 cases, similar reactions were also reported in the mothers), and reactions to cephalosporins were recorded in 2 patients. Life-threatening ADRs (DRESS syndrome) were observed in 2 patients receiving vancomycin; one of these patients subsequently died after hospitalization due to cardiac complications associated with the ADR. Both patients carried the *HLA-A*32:01* risk allele. These pharmacogenetic associations represented the only findings in our cohort supported by high-level evidence (ClinPGx/PharmGKB Level 1A–1B).

Among “Variant Annotations” entries in ClinPGx/PharmGKB, which represent lower-evidence associations, explanations were identified for three additional cases: *CYP2D6*4*-associated poor metabolism resulting in extrapyramidal symptoms during metoclopramide treatment, the association of *HLA-B*15:01* with cutaneous ADRs during amoxicillin therapy, and increased susceptibility to hypersensitivity reactions to penicillin antibiotics among *HLA-A*02:01* carriers.

For the remaining documented ADRs (including post-vaccination allergic reactions, bronchospasm associated with lidocaine, and reactions to miramistin, reamberin, B-group vitamins, diphenhydramine, and novocaine), corresponding PGx associations were absent from the ClinPGx/PharmGKB databases. These reactions most likely represent classical idiosyncratic or immune-mediated ADRs, in which genetic factors currently characterized within pharmacogenomic databases do not play a predominant role. The absence of PGx explanations for such reactions highlights that not all ADRs can be prevented through currently available pharmacogenomic testing, and their prevention requires additional strategies, including detailed allergy history assessment, skin testing, and alternative approaches for managing patients with multiple drug intolerance [63].

Polypharmacy (>5 drugs per hospitalization) was observed in 55% of patients; 29% of patients received more than three drugs from the same therapeutic class, most commonly antibiotics (13% of patients, with up to 11 antibiotics prescribed during a single hospitalization). The highest level of polypharmacy was observed within the antibiotic category, reflecting the complexity of empirical treatment selection in patients with severe infectious complications, antimicrobial resistance, and frequent changes in therapeutic regimens.

The present analysis has several limitations. First, information regarding medication history and ADRs was retrospectively extracted from medical records, which may have resulted in incomplete data, particularly regarding assessment of causal relationships between drug exposure and clinical deterioration. Second, medical documentation often lacked differentiation between allergic reactions (rash, urticaria, edema) and non-allergic toxic effects (lethargy, vomiting, dysarthria, extrapyramidal symptoms, respiratory depression), potentially affecting the interpretation of ADR frequency and structure. Third, pharmacogenetic testing was not performed in all patients; therefore, the actual frequency of PGx-associated ADRs may be underestimated due to undetected cases. Fourth, a substantial proportion of patients had severe monogenic disorders with progressive disease courses, complicating the assessment of treatment efficacy because clinical deterioration could reflect the natural progression of the underlying disease rather than drug inefficacy. Fifth, the completeness of medical documentation varied considerably, ranging from initial departmental assessments to comprehensive discharge summaries containing detailed medical histories spanning several years of life. Sixth, the data were obtained from different clinical departments and healthcare institutions in Moscow.

As a clinical example, we identified a patient from our cohort with genetically confirmed glucose-6-phosphate dehydrogenase (G6PD) deficiency (variant *G6PD*(NM_001042351.3):c.1178G>A (p.Arg393His)) accompanied by Gilbert syndrome. The primary disorder manifested as chronic hemolysis with episodes of hemolytic crises, one of which was associated with a decrease in hemoglobin concentration to 47 g/L and required blood transfusion. Examination revealed a marked reduction in G6PD activity (20 mU/10^9^ erythrocytes; reference range 244–299), reticulocytosis (15.5%), hyperbilirubinemia up to 81 μmol/L, moderate splenomegaly, and signs of iron overload (ferritin up to 545 ng/mL). The patient received therapy with folic acid, ursodeoxycholic acid, deferasirox, and cholecalciferol. Based on pharmacogenomic analysis, an individualized list of medications and other factors capable of inducing hemolysis in the context of G6PD deficiency was generated. This information enabled personalization of subsequent recommendations for pharmacotherapy and prevention of hemolytic crises.

## 8. Recommendations for the Clinical Use of Pharmacogenomics in Pediatrics

Table 2 summarizes the key components required to improve the clinical applicability of pharmacogenomic testing in children. The proposed approach includes mandatory consideration of patient age, ontogenetic changes in enzyme and transporter activity, the level of evidence supporting recommendations, limitations of sequencing technologies, and the need for multidisciplinary clinical interpretation.

**Table 2.** Limitations of current pharmacogenomic reports (including those used in the Russian Federation) and recommended requirements for pediatric PGx interpretation.

| Report component | Current status | Recommended approach for pediatric PGx interpretation | Rationale |
| --- | --- | --- | --- |
| Patient age | Frequently absent or not considered during report generation | Mandatory inclusion of patient age and developmental stage | Activity of drug-metabolizing enzymes and transport systems changes throughout ontogeny |
| Ontogenetic correction | Usually absent; recommendations are based on adult-derived algorithms | Specify age-related limitations of recommendation applicability (e.g., “this recommendation has not been validated in children younger than X years”) | Particularly important for genes with age-dependent expression patterns: CYP3A4/3A7, CYP2C19, CYP2D6, UGT1A1 |
| Pediatric applicability | Pediatric data are rarely reported separately | Include a dedicated annotation indicating “pediatric applicability” | Most PGx recommendations have been developed based on adult cohorts |
| Evidence level | Evidence categories may be absent or mixed with exploratory research findings | Report evidence level according to CPIC/DPWG/FDA; classify recommendations lacking sufficient clinical validation as informational only | Prevents overinterpretation of uncertain findings and reduces the risk of iatrogenic harm |
| Dosing recommendations | Adult dosing algorithms are often applied directly | Specify the need for age-, weight-, or ontogeny-adjusted correction; indicate cases where adult recommendations are extrapolated | Pediatric pharmacokinetics is determined by body weight, organ maturation, and development of metabolic pathways |
| CNV *CYP2D6* | Not assessed in most WES-based approaches | Explicitly state the limitation of *CYP2D6* interpretation without copy-number analysis | *CYP2D6* duplications and deletions may result in incorrect metabolizer phenotype classification |
| Responsible specialist | No standardized procedure exists for PGx result interpretation | Joint interpretation by a clinical pharmacologist and a medical geneticist | PGx requires integration of genotype, phenotype, drug therapy, and clinical context |

Analysis of pharmacogenomic recommendations demonstrated that the application of adult-derived algorithms in children is limited by age-dependent changes in the expression of drug-metabolizing enzymes and transport proteins (Table 1). The most substantial limitations were identified for genes with dynamic ontogenetic regulation of expression, including *CYP3A4/3A7*, *CYP2C19*, *CYP2D6*, and *UGT1A1*, for which the same genotype may correspond to different functional phenotypes depending on the patient’s age. For genes with relatively stable expression patterns (*TPMT*, *NUDT15*, *CYP3A5*), the application of existing recommendations requires fewer adjustments. Based on these findings, we developed a model of age-oriented interpretation of PGx data that integrates the interaction between genotype, age, and developmental biology (Figure 6A).

**Figure 6.**
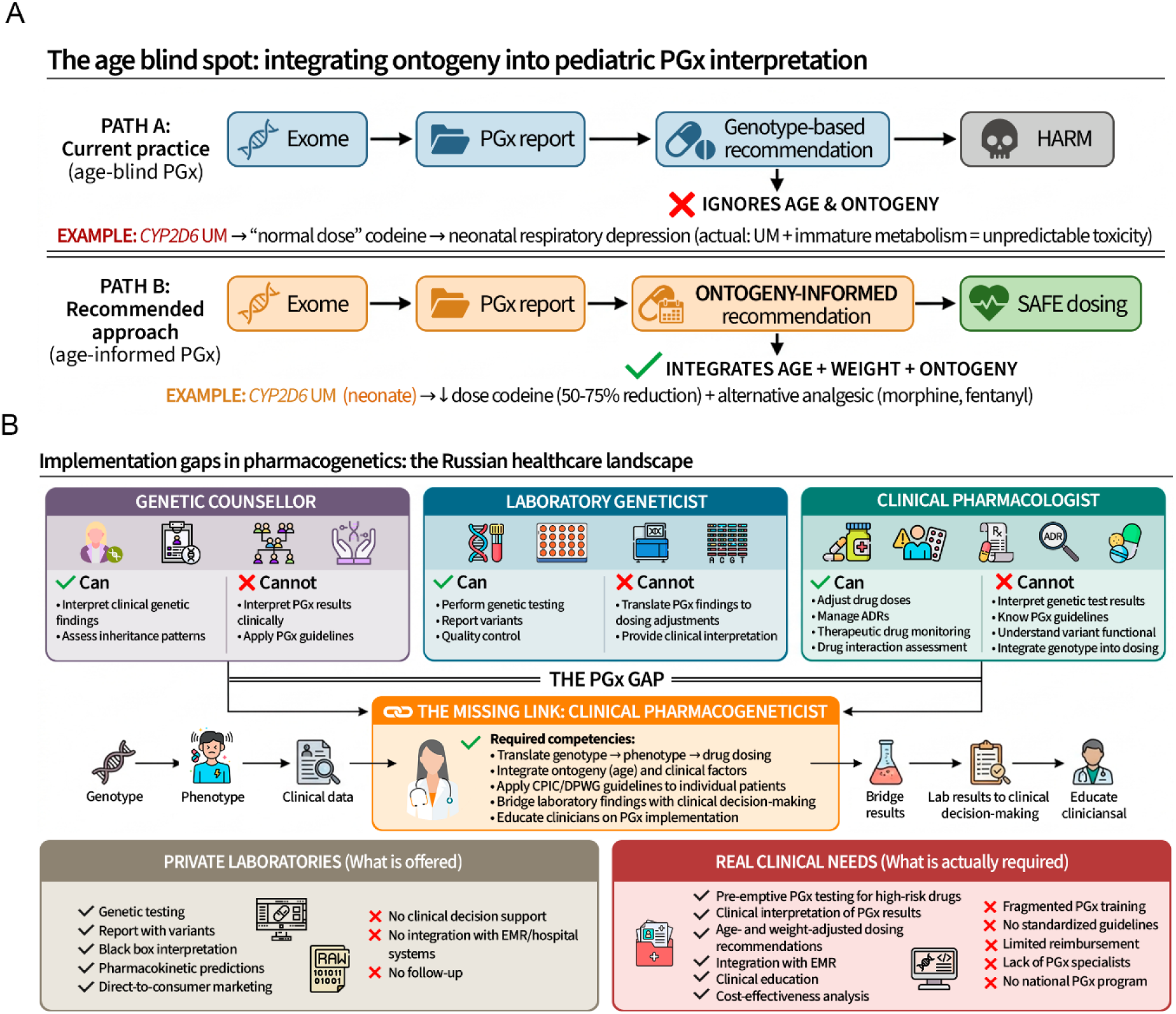
Current state of pharmacogenomics implementation in clinical practice in the Russian Federation. (А) The “age blind spot” in pharmacogenomics: integration of ontogeny into pediatric PGx interpretation. Blue boxes represent current practice (without consideration of age), potentially leading to inappropriate clinical decisions. Orange boxes represent the recommended approach incorporating ontogenetic information. (B) Barriers to implementation of clinical pharmacogenomics within the Russian healthcare system.

Evaluation of the current model of pharmacogenomic testing revealed the absence of a unified clinical interpretation workflow that ensures the transition from identification of a genetic variant to individualized drug therapy. At present, separate stages of the PGx workflow are distributed among different specialists: a medical geneticist provides expertise in interpretation of genetic data and assessment of hereditary factors, a laboratory geneticist performs testing and analytical quality control, and a clinical pharmacologist is responsible for rational medication use. However, there is no unified component integrating genetic information, pharmacological recommendations, patient age-related characteristics, and clinical status.

An additional limitation is insufficient standardization of pharmacogenomic report structure. In particular, existing reports may include genetic variants without assessment of evidence level, age-specific applicability, or clinical relevance, limiting the safe implementation of results in pediatric practice. Based on the identified limitations, we propose a clinical implementation model for PGx that includes multidisciplinary interpretation of results, application of evidence-based recommendations, and integration of pharmacogenomic data into clinical decision support systems (Figure 6B).

## 9. Conclusions

A child is not a smaller version of an adult patient, and the present study demonstrates this principle using quantitative data. After filtering for clinical relevance (evidence level 1A–2B and pediatric applicability), only 5% of initial pharmacogenomic annotations remained, with the majority corresponding to alleles with altered function. Comparison of these genotype-based recommendations with the actual ontogenetic status of enzymes in newborns demonstrated concordance in less than half of the analyzed gene–drug pairs.

Retrospective analysis of 100 medical records demonstrated the clinical consequences of this gap: despite an ADR frequency of 21%, only 2% of observed cases could be explained by high-evidence pharmacogenomic annotations, indicating a substantial discrepancy between real-world clinical practice and the coverage provided by current pharmacogenomic databases.

Using the relative population burden metric (allele frequency × ADR episode cost), we prioritized 27 gene–variant–drug–ADR combinations and identified associations potentially suitable for clinical implementation, including *UGT1A1*28*–irinotecan-induced neutropenia and *HLA-A*31:01*–carbamazepine-induced severe cutaneous ADR. However, this metric is a preliminary prioritization tool and does not account for variant penetrance or baseline ADR incidence. Similarly, the estimates obtained using the three-component model are based on published epidemiological data and clinical guidelines and should be interpreted as population-level approximations rather than precise measures. Finally, the presented clinical case of a patient with glucose-6-phosphate dehydrogenase deficiency and Gilbert syndrome illustrates the practical value of personalized pharmacogenomic counseling in individuals with combined genetic disorders.

To improve the clinical applicability of pharmacogenomic results, implementation of age-oriented reports is required, with explicit indication of recommendation applicability within specific age ranges, a minimum evidence standard based on CPIC/DPWG/FDA criteria, and mandatory multidisciplinary interaction between medical geneticists, clinical pharmacologists, and laboratory specialists, particularly because a separate medical specialty of “pharmacogeneticist” does not currently exist in Russia.

Future studies should focus on expanding the ontogenetic evidence base for genes with limited developmental data (*NUDT15*, *VKORC1*) and on prospective validation of the proposed framework for prioritization and interpretation of pharmacogenomic variants in real-world pediatric clinical practice.

## Supporting information

All parameters were extracted from published sources (Table S1).

pairs for which cost data were available (Figure 3; Table S2).

## Additional Files

Table S1. Estimated pediatric utilization of pharmacogenomically relevant drugs in the Russian population (0–18 years). Table S2. Integrated pharmacogenomic dataset containing allele frequencies, ClinPGx evidence, adverse drug reaction (ADR) cost estimates, and calculated relative burden for all “gene–variant–drug–ADR” combinations.

## Institutional Review Board Statement

This study was conducted in accordance with the Declaration of Helsinki, and approval was obtained from the Local Research Ethics Committee of Russian National Medical University (Protocol No. 241, from 26 June 2024), and all participants provided written informed consent prior to data collection.

## Author Contributions

Buianova A.A. – formal analysis, conceptualization, methodology, visualization, software, writing–original draft preparation; Cheranev V.V., Kuznetsov M.Iu. – data curation, software; Belova V.A. – methodology, investigation; Repinskaia Z.A. – software, investigation.

## Informed Consent Statement

Informed consent was obtained from all subjects involved in this study.

## Data Availability Statement

The individual-level DNA sequencing data generated in this study constitute sensitive human genomic data and are subject to legal and ethical restrictions under national regulations governing human genetic data protection in the Russian Federation. Access to individual-level data may be granted upon reasonable request to the corresponding author, subject to approval by the relevant institutional review board and in compliance with applicable legal and ethical requirements.

